# Semaglutide Initiation and Treatment Duration On Suicidality Risk in US Veterans With Type 2 Diabetes

**DOI:** 10.64898/2026.04.17.26351118

**Authors:** Ana I. Maldonado, Kent R. Heberer, Julie A. Lynch, Steven Cogill, Shriram Nallamshetty, Ying Q. Chen, Mei-Chung Shih, Adam P. Bress, Jennifer S. Lee

**Affiliations:** VA Palo Alto Healthcare System, Palo Alto, California, USA; VA Palo Alto Cooperative Studies Program Coordinating Center, Palo Alto, California, USA; VA Salt Lake City Healthcare System, Salt Lake City, Utah, USA; Division of Epidemiology, School of Medicine, University of Utah, Salt Lake City, Utah, USA; Division of Endocrinology, Gerontology, and Metabolism, Department of Medicine, and by courtesy, Department of Epidemiology and Population Health, Stanford University School of Medicine, Stanford, CA, USA; Division of Cardiovascular Medicine, Stanford School of Medicine, Stanford, California, USA; Stanford Prevention Research Center, Department of Medicine, Stanford University, Stanford, California, USA; Department of Biomedical Data Science, Stanford University School of Medicine, Stanford, California, United States of America; Department of Population Health Sciences, School of Medicine, University of Utah, Salt Lake City, Utah, USA

## Abstract

**Importance:** Semaglutide, a glucagon-like peptide-1 receptor agonist (GLP-1RA), is a highly effective medication to treat type 2 diabetes and obesity. However, concerns about potential suicidality persist, creating clinical uncertainty about its neuropsychiatric safety.

**Objective:** To assess risks of suicidality after initiating semaglutide compared to initiating SGLT2i and by duration of continuous semaglutide treatment.

**Design:** Active-comparator, new-user target trial emulation to estimate inverse probability–weighted marginal cause-specific hazard ratios (HRs). For duration-of-treatment analyses, we used clone–censor–weight methods to estimate exposure-adjusted effects.

**Setting:** Veterans Health Administration.

**Participants:** U.S. Veterans with type 2 diabetes receiving care from March 1, 2018 to September 1, 2025.

**Exposure:** Initiation of semaglutide vs SGLT2i; duration of semaglutide use (≤6, 7–12, >12 months).

**Outcomes:** Incident suicidal ideation; suicide attempt or death; and a composite outcome.

**Results:** A total of 102,361 Veterans met inclusion criteria, including 11,478 new initiators of semaglutide and 90,883 new initiators of an SGLT2i. After overlap weighting, baseline characteristics were well balanced between treatment groups (mean [SD] age, 60.1 [11.7] years; BMI, 37.8 [6.8] kg/m^2^; hemoglobin A1c, 7.0% [1.4]; 85.5% male; 61.9% non-Hispanic White). During a median follow-up of 2.2 years, 9077 incident suicidal ideation events and 696 suicide attempts or deaths occurred. The incidence rate of suicidal ideation was 56.3 and 37.7 per 1000 person-years among semaglutide initiators and SGLT2i initiators, respectively (hazard ratio [HR], 0.99; 95% CI, 0.93-1.06; P = 0.86). For suicide attempts or deaths, the incidence rates were 4.30 and 2.64 per 1000 person-years, respectively (HR, 1.05; 95% CI, 0.84-1.31; P = .86). In adherence-adjusted analyses, sustained semaglutide treatment for more than 12 months, compared with 6 or fewer months, was associated with a 74% lower risk of suicide attempts or deaths (HR, 0.27; 95% CI, 0.14-0.54; P<.001).

**Conclusion:** Among U.S. Veterans with type 2 diabetes, initiators of semaglutide were not observed to have an increased risk of suicidality compared with initiators of SGLT2i. Those with longer semaglutide treatment (beyond 12 months) had decreased risk of suicide attempt or death, suggesting longer term treatment is safe and may protect against for those outcomes.

## Introduction

Semaglutide, a glucagon-like peptide-1 receptor agonist (GLP-1RA), is among the most widely prescribed therapies for type 2 diabetes (T2D) and obesity, with an estimated 15 million adults in the United States (U.S.) currently using the medication.^1^ Although semaglutide is highly effective for glycemic control, weight reduction, and cardiovascular risk reduction, concerns have emerged regarding potential associations with suicidality. Semaglutide approved for weight management carries a boxed warning related to suicidal behavior and ideation based on clinical trial findings.^2^

Recent meta-analyses of randomized control trials (RCTs), observational cohort studies, and pharmacovigilance studies have not found evidence that semaglutide increases risk of suicidality.^3–6^ However, uncertainty remains regarding whether risk varies by duration of GLA-1RA exposure. Pharmacovigilance analyses have suggested that adverse psychiatric events may cluster shortly after treatment initiation,^6,7^ raising concern for an acute vulnerability period during dose escalation. In contrast, some observational studies reporting lower rates of suicidality among users of semaglutide^8^ may reflect longer-term exposure among individuals who tolerate and remain adherent to treatment. Few studies have rigorously evaluated whether suicidality risk is confined to an early exposure window or changes with sustained treatment. Prior investigations have been limited by reliance on intention-to-treat (ITT) designs that do not account for treatment discontinuation or non-adherence, heterogeneous data sources, small numbers of suicide events, and relatively short follow-up. Moreover, limited evidence exists in U.S. Veterans, a population with elevated risk of suicidal ideation, attempts, and suicide mortality.^9^

To address these gaps, we emulated a target trial in a nationwide cohort of Veterans with T2D receiving care in the Veterans Health Administration. Using pharmacy dispensing data, longitudinal clinical covariates, and extended follow-up, we evaluated the risk of incident suicidal ideation and suicide attempts or death associated with semaglutide initiation compared with sodium-glucose cotransporter-2 inhibitor (SGLT2i) initiation. We additionally applied clone–censor–weighting (CCW) methods to examine how duration of semaglutide treatment (≤6 months, 7–12 months, and >12 months) was associated with suicidality risk.

## Methods

### Study Design

We emulated two target trials to estimate the risk of suicidal ideation and attempts/completion in a retrospective nationwide cohort of US Veterans with T2D. First, an active-comparator new user design with propensity score (PS) weighting and an ITT strategy was used to estimate the average treatment effect of the overlap population among initiators of semaglutide compared to initiators of SGLT2i. Second, we used CCW to simulate the effect of duration of continuous exposure among initiators of semaglutide on the risk of suicidality outcomes across three treatment arms: short term (≤6 months), intermediate (7-12 months), and long-term (>12 months).

### Participants and Exposure

Veterans aged ≥18 years with T2D and a prescription for metformin who initiated semaglutide or an SGLT2i between March 1, 2018, and March 1, 2025 were identified for inclusion. The index date was the first outpatient pharmacy fill of either drug. Eligibility required ≥1 diagnosis code for T2D within two years before index and a metformin fill within 90 days. Exclusions were prior type 1 or secondary diabetes, exposure to other antidiabetic drugs, or >30 days of insulin use within 10 years. The treatment strategies were initiation of semaglutide vs SGLT2i, identified using pharmacy dispensing data. Veterans who initiated both simultaneously or neither were excluded. See Supplement for patient selection flow diagram and criteria.

### Follow-up and Outcomes

Follow-up began on index date and continued until a primary suicide outcome (i.e. suicidality), death, or end of study follow-up (September 1, 2025). The primary suicide outcomes were 1) incident suicidal ideation, defined using a validated algorithm,^10^ which includes diagnosis codes, screening positive on either clinician-administered questionnaires or self-report survey questionnaires, 2) incident suicide attempts/completions from clinician-administered risk assessments, and 3) composite outcome of the first event to occur between suicidal ideation and suicidal attempt/completion. Each primary suicide outcome was assessed independently.

### Baseline Covariates

Baseline covariates were selected a priori for their potential to confound the association between semaglutide and SGLT2i initiation and suicide risk, including mental health conditions, mental health treatment (pharmacotherapy and psychotherapy), and history of suicide ideation or attempts (Table S2). Baseline covariates were assessed during the two years before the index date using validated algorithms, except history of suicidality which was assessed as any exposure prior to index date.

### Statistical Analysis

To address measured confounding by baseline covariates, we estimated PS for semaglutide initiation using logistic regression with all baseline covariates included in the model^11^ and implemented them using overlap weighting with the PSweight R package.^12^ Baseline covariate balance was assessed before and after weighting using absolute standardized mean differences, with values <0.01 considered indicative of adequate balance. Missing baseline covariates with <20% missingness and judged to be missing at random were imputed using multiple imputation with chained equations using the mice R Package,^13^ generating 10 imputed datasets.

The first target trial emulation followed the ITT principle. For each primary suicide outcome, we calculated the unweighted cumulative incidence (number of events divided by persons at risk) and incidence rates (number of events divided by person-time at risk) by treatment group. We estimated cumulative incidence functions for primary suicide outcomes by treatment group using Kaplan Meier methods and compared groups with the log rank test. Kaplan Meier curves are presented for unweighted and overlap weighted cohorts. Adjusted hazard ratios (HRs) and 95% confidence intervals (CIs) were estimated using Cox proportional hazards models that incorporated overlap weighting based on PS. The set of covariates included in the PS model, and therefore in adjusted HR estimates, was identical to the baseline covariates listed above, and was implemented with the PSweight package.

To assess semaglutide exposure duration effects on risk of primary suicide outcomes, we implemented per-protocol methods using a CCW scheme^14,15^ with the sub-cohort of semaglutide initiators. Semaglutide exposure was assessed over 28-day periods starting with index initiation, with continuing exposure defined as 14 or more days of coverage during the 28-day period. For a detailed description of CCW methods, see the Supplement.

The conditional HR for suicide attempt/completion and for suicidal ideation was estimated using pooled logistic regression with a quasi-binomial distribution using the treatment arm and period as predictors and weighted by unstabilized IPAW/IPCW. Statistical significance of the coefficients of the fitted model were assessed using R package lmtest^16^ and R package sandwich for the covariance estimates.^17,18^ Average event risk curves were estimated by simulating the incidence of cases if all patients had adhered to each of the three treatment arms. The 95% confidence bands were generated by using bootstrap resampling methods for 200 iterations.

## Results

### Study Population

Among 814,019 Veterans who initiated semaglutide or an SGLT2i between March 1, 2018, and March 1, 2025, a total of 102,361 met study eligibility criteria (Figure S1). Among these, 11,478 initiated semaglutide and 90,883 initiated an SGLT2i. Nearly all SGLT2i initiators received empagliflozin (n=90,816). The remainder (n=67) received dapagliflozin or canagliflozin.

### Baseline Characteristics

At baseline, semaglutide users were more likely than SGLT2i users to be female (17.2% vs 6.3%; p<.001) and Black (21.6% vs 17.8%; p<.001). Overall, semaglutide users were also younger (mean [SD] age, 59.2 [11.7] years vs 64.6 [11.9] years), had a higher body mass index (38.8 [7.4] vs 33.6 [6.4]), and had a lower hemoglobin A1c (7.0% [1.4] vs 7.3% [1.5]). After application of overlap weighting, all covariates were well-balanced, with standardized mean differences <0.1 (Table 1).

**Table 1.** Baseline Covariates Between Initiators of Semaglutide and an SGLT2i Before and After Overlap Weighting^a^.

| Characteristics<sup>b</sup> | Before | Weighting | SMD | After | Weighting | SMD<sup>d</sup> |
| --- | --- | --- | --- | --- | --- | --- |
|  | Semaglutide,<br>No. (%)<br>(n=11478) | SGLT2i,<br>No. (%)<br>(n=90883) |  | Semaglutide,<br>No. (%) | SGLT2i,<br>No. (%) |  |
| History of Suicide |  |  |  |  |  |  |
| Attempt | 501 (4.4%) | 2451 (2.7%) | 0.09 | 3.9% | 3.9% | 2.4 x10<sup>-12</sup> |
| Ideation | 2832 (24.7%) | 16088 (17.7%) | 0.17 | 23.1% | 23.1% | 3.0 x10<sup>-12</sup> |
| Currently Treated for Mental Health Concerns | 3707 (32.3%) | 18590 (20.5%) | 0.27 | 29.6% | 29.6% | 4.2 x10<sup>-13</sup> |
| Age, mean (SD), y | 59.2 (11.7) | 64.6 (11.9) | 0.46 | 60.1 (11.7) | 60.1 (12.2) | 1.0 x10<sup>-12</sup> |
| Sex |  |  |  |  |  |  |
| Female | 1978 (17.2%) | 5749 (6.3%) | 0.34 | 14.5% | 14.5% | 1.0 x10<sup>-12</sup> |
| Male | 9500 (82.8%) | 85134 (93.7%) | NA | 85.5% | 85.5% | NA |
| Race/Ethnicity (ref: Non-Hispanic White) |  |  |  |  |  |  |
| American Indian or Alaska Native | 66 (0.6%) | 641 (0.7%) | 0.02 | 0.6% | 0.6% | 1.8 x10<sup>-12</sup> |
| Asian | 132 (1.2%) | 1600 (1.8%) | 0.05 | 1.2% | 1.2% | 1.9 x10<sup>-12</sup> |
| Black or African American | 2480 (21.6%) | 16218 (17.8%) | 0.09 | 20.6% | 20.6% | 1.9 x10<sup>-12</sup> |
| Hispanic or Latino | 948 (8.3%) | 6951 (7.6%) | 0.02 | 8.1% | 8.1% | 2.7 x10<sup>-12</sup> |
| Hawaiian or Pacific Islander | 131 (1.1%) | 979 (1.1%) | 0.01 | 1.1% | 1.1% | 3.4 x10<sup>-13</sup> |
| Non-Hispanic White | 6992 (60.9%) | 58818 (64.7%) | NA | 61.9% | 61.9% | NA |
| Other or Not Reported | 729 (6.4%) | 5676 (6.2%) | 0.00 | 6.4% | 6.4% | 5.5 x10<sup>-13</sup> |
| Index Year (ref: 2024) |  |  |  |  |  |  |
| 2018 | 21 (0.2%) | 887 (1.0%) | 0.10 | 0.2% | 0.2% | 3.8 x10<sup>-10</sup> |
| 2019 | 192 (1.7%) | 2592 (2.9%) | 0.08 | 1.9% | 1.9% | 2.5 x10<sup>-12</sup> |
| 2020 | 321 (2.8%) | 4677 (5.1%) | 0.12 | 3.2% | 3.2% | 3.2 x10<sup>-12</sup> |
| 2021 | 1190 (10.4%) | 10746 (11.8%) | 0.05 | 10.8% | 10.8% | 6.2 x10<sup>-12</sup> |
| 2022 | 1960 (17.1%) | 17915 (19.7%) | 0.07 | 17.5% | 17.5% | 8.3 x10<sup>-12</sup> |
| 2023 | 3177 (27.7%) | 23864 (26.3%) | 0.03 | 27.5% | 27.5% | 1.1 x10<sup>-11</sup> |
| 2024 | 4167 (36.3%) | 26139 (28.8%) | NA | 34.9% | 34.9% | NA |
| 2025 | 450 (3.9%) | 4063 (4.5%) | 0.03 | 4.0% | 4.0% | 3.7 x10<sup>-12</sup> |
| Clinical and laboratory measurements, mean (SD) |  |  |  |  |  |  |
| Body Mass Index (kg/m<sup>2</sup>) | 38.8 (7.4) | 33.6 (6.4) | 0.74 | 37.8 (6.8) | 37.8 (7.8) | 5.4 x10<sup>-14</sup> |
| Systolic Blood Pressure, mmHg | 133.7 (15.8) | 135.1 (17.0) | 0.09 | 133.9 (15.9) | 133.9 (16.0) | 2.4 x10<sup>-12</sup> |
| Total Cholesterol, mg/dL | 167.7 (47.0) | 164.0 (48.1) | 0.08 | 166.8 (46.8) | 166.8 (45.6) | 7.2 x10<sup>-13</sup> |
| Triglyceride, mg/dL | 188.5 (164.1) | 195.8 (175.5) | 0.04 | 192.5 (162.7) | 192.5 (154.0) | 2.2 x10<sup>-12</sup> |
| High-Density Lipoprotein, mg/dL | 41.8 (11.4) | 41.3 (11.3) | 0.04 | 41.4 (11.2) | 41.4 (11.4) | 2.2 x10<sup>-12</sup> |
| Low-Density Lipoprotein, mg/dL | 94.1 (38.6) | 89.9 (38.9) | 0.11 | 93.1 (38.7) | 93.1 (38.3) | 1.6 x10<sup>-12</sup> |
| Hemoglobin A1c, % | 7.0 (1.4) | 7.3 (1.5) | 0.23 | 7.0 (1.4) | 7.0 (1.2) | 6.6 x10<sup>-13</sup> |
| Lifestyle Factors |  |  |  |  |  |  |
| AUDIT-C <sup>c</sup> | 1.5 (2.1) | 1.7 (2.5) | 0.08 | 1.5 (2.2) | 1.5 (2.3) | 1.7x10<sup>-12</sup> |
| Smoking History (Yes) | 5797 (50.5%) | 49827 (54.8%) | 0.09 | 51.0% | 51.0% | 7.1x10<sup>-14</sup> |
| Charlson Comorbidity Index |  |  |  |  |  |  |
| Cerebrovascular Disease | 618 (5.4%) | 6702 (7.4%) | 0.08 | 5.7% | 5.7% | 4.2 x10<sup>-13</sup> |
| Congestive Heart Failure | 807 (7.0%) | 12882 (14.2%) | 0.23 | 7.7% | 7.7% | 4.4 x10<sup>-13</sup> |
| Chronic Pulmonary Disease | 2219 (19.3%) | 17010 (18.7%) | 0.02 | 19.0% | 19.0% | 9.1 x10<sup>-13</sup> |
| Dementia | 210 (1.8%) | 2306 (2.5%) | 0.05 | 1.9% | 1.9% | 1.2 x10<sup>-13</sup> |
| Diabetes with Complications | 3591 (31.3%) | 32079 (35.3%) | 0.09 | 32.2% | 32.2% | 3.0 x10<sup>-12</sup> |
| Hemiplegia or Paraplegia | 90 (0.8%) | 435 (0.5%) | 0.04 | 0.7% | 0.7% | 4.1 x10<sup>-13</sup> |
| **HIV or AIDS** | 87 (0.8%) | 377 (0.4%) | 0.04 | 0.7% | 0.7% | 1.9 x10<sup>-13</sup> |
| **Liver Disease (Mild)** | 1802 (15.7%) | 8954 (9.9%) | 0.18 | 14.5% | 14.5% | 3.1 x10<sup>-12</sup> |
| **Liver Disease (Severe)** | 146 (1.3%) | 1192 (1.3%) | 0.00 | 1.3% | 1.3% | 2.3 x10<sup>-13</sup> |
| **Malignancy** | 1576 (13.7%) | 13724 (15.1%) | 0.04 | 13.8% | 13.8% | 1.3 x10<sup>-12</sup> |
| **Metastasis** | 67 (0.6%) | 557 (0.6%) | 0.00 | 0.6% | 0.6% | 8.5 x10<sup>-13</sup> |
| **Myocardial Infarction** | 305 (2.7%) | 4221 (4.6%) | 0.11 | 2.9% | 2.9% | 1.1 x10<sup>-12</sup> |
| **Peptic Ulcer** | 50 (0.4%) | 591 (0.7%) | 0.03 | 0.5% | 0.5% | 8.9 x10<sup>-13</sup> |
| **Peripheral Vascular Disease** | 713 (6.2%) | 7810 (8.6%) | 0.09 | 6.5% | 6.5% | 2.3 x10<sup>-13</sup> |
| **Renal Disease** | 880 (7.7%) | 9004 (9.9%) | 0.08 | 7.9% | 7.9% | 2.4 x10<sup>-12</sup> |
| **Rheumatic Disorders** | 209 (1.8%) | 1325 (1.5%) | 0.03 | 1.7% | 1.7% | 1.0 x10<sup>-13</sup> |
| **Mental Health Risk Factors** |  |  |  |  |  |  |
| Adjustment and Reaction to Stress | 1407 (12.3%) | 7921 (8.7%) | 0.12 | 11.5% | 11.5% | 7.2 x10<sup>-13</sup> |
| PTSD | 3533 (30.8%) | 19815 (21.8%) | 0.21 | 29.0% | 29.0% | 2.0 x10<sup>-12</sup> |
| Psychotic Disorders | 235 (2.0%) | 1378 (1.5%) | 0.04 | 1.9% | 1.9% | 7.0 x10<sup>-13</sup> |
| Anxiety Disorders | 3128 (27.3%) | 16607 (18.3%) | 0.22 | 25.4% | 25.4% | 2.4 x10<sup>-12</sup> |
| Bipolar Depression Disorders | 484 (4.2%) | 2136 (2.4%) | 0.10 | 3.7% | 3.7% | 1.4 x10<sup>-12</sup> |
| Substance Use Disorders | 2069 (18.0%) | 16008 (17.6%) | 0.01 | 17.8% | 17.8% | 7.2 x10<sup>-13</sup> |
| Personality Disorders | 299 (2.6%) | 1114 (1.2%) | 0.10 | 2.2% | 2.2% | 4.3 x10<sup>-13</sup> |
| Other Mood Disorders | 591 (5.1%) | 3150 (3.5%) | 0.08 | 4.7% | 4.7% | 8.2 x10<sup>-13</sup> |
| Lifestyle Problems | 1498 (13.1%) | 12202 (13.4%) | 0.01 | 13.0% | 13.0% | 1.4 x10<sup>-12</sup> |
| Adverse Socioeconomic and Psychosocial Problems | 2527 (22.0%) | 16412 (18.1%) | 0.10 | 20.8% | 20.8% | 1.5 x10<sup>-12</sup> |
| Depression Disorders | 4447 (38.7%) | 24199 (26.6%) | 0.26 | 36.2% | 36.2% | 2.1 x10<sup>-12</sup> |
| Trauma History | 82 (0.7%) | 325 (0.4%) | 0.05 | 0.6% | 0.6% | 1.4 x10<sup>-13</sup> |
| Family Mental Illness History | 43 (0.4%) | 196 (0.2%) | 0.03 | 0.3% | 0.3% | 6.8 x10<sup>-13</sup> |
| Behavioral Disorders including Eating and Sleep | 1124 (9.8%) | 6218 (6.8%) | 0.11 | 9.0% | 9.0% | 1.2 x10<sup>-12</sup> |
| **Concurrent Medications** |  |  |  |  |  |  |
| Antidepressants | 5759 (50.2%) | 33063 (36.4%) | 0.28 | 47.6% | 47.6% | 3.6 x10<sup>-13</sup> |
| Antiepileptics | 1491 (13.0%) | 6476 (7.1%) | 0.20 | 11.4% | 11.4% | 2.1 x10<sup>-13</sup> |
| Hypnotics and Sedatives | 1585 (13.8%) | 9030 (9.9%) | 0.12 | 12.9% | 12.9% | 2.5 x10<sup>-13</sup> |
| Antipsychotics | 1119 (9.7%) | 5483 (6.0%) | 0.14 | 8.8% | 8.8% | 8.2 x10<sup>-13</sup> |

**Abbreviations: SGLT2i: Sodium-Glucose Co-transporter inhibitor; SMD: Standardized Mean Difference; AUDIT-C: Alcohol Use Disorders Identification Test – Consumption**
<sup>a</sup>Data for before weighting are presented as number (percentage) of participants, and data for after weighting as percentage of participants unless otherwise indicated. The total numbers of patients in the post-overlap weighted columns were omitted because the numbers were slightly different as a result of the weighting.
<sup>b</sup>Values were imputed for the following (Semaglutide/SGLT2i): BMI = 454/4222, total cholesterol = 952/6989, triglycerides = 1047/7945, HDL = 962/7099, LDL = 1048/7863, blood pressure = 387/3122, HbA1c = 307/2373, AUDIT-C = 131/1387
<sup>c</sup>AUDIT-C raw scores range from 0-12, with 0 indicating no drinking behavior.
<sup>d</sup>All values less than 0.01

**Table 2.** Absolute Incidence Rates and Hazard Ratios of Suicidality Outcomes in Unweighted and Overlap Weighted Cohort.

|  | Events | At-Risk Time<br>(years) | Incidence Rate<br>(95% CI) per 1000<br>Person-Years | Unweighted |  | Overlap Weighting |  |
| --- | --- | --- | --- | --- | --- | --- | --- |
|  |  |  |  | HR<sup>1</sup> (95% CI) | p-value | HR<sup>1</sup> (95% CI) | p-value |
| Overall Cohort |  |  |  |  |  |  |  |
| Suicide Ideation |  |  |  |  |  |  |  |
| Semaglutide Initiators | 1321 | 23455 | 56.3 (53.4, 59.3) | 1.47 (1.39, 1.56) | P < .001* | 0.99 (0.93, 1.06) | P=0.86 |
| SGLT2i Initiators | 7756 | 210477 | 36.8 (36.0, 37.7) |  |  |  |  |
| Suicide Attempt/Completion |  |  |  |  |  |  |  |
| Semaglutide Initiators | 109 | 25358 | 4.30 (3.53, 5.18) | 1.61 (1.31, 1.98) | P<.001* | 1.05 (0.84, 1.31) | P=0.68 |
| SGLT2i Initiators | 587 | 222634 | 2.64 (2.43, 2.86) |  |  |  |  |
| Without Prior History of Suicidality |  |  |  |  |  |  |  |
| Suicide Ideation |  |  |  |  |  |  |  |
| Semaglutide Initiators | 473 | 18774 | 25.2 (23.0, 27.5) | 1.36 (1.24, 1.50) | P<.001* | 1.00 (0.90, 1.11) | P=0.98 |
| SGLT2i Initiators | 3304 | 179975 | 18.4 (17.7, 19.0) |  |  |  |  |
| Suicide Attempt/Completion |  |  |  |  |  |  |  |
| Semaglutide Initiators | 22 | 19416 | 1.13 (0.71, 1.71) | 1.17 (0.75, 1.82) | P=.49 | 0.99 (0.62, 1.57) | P=0.95 |
| SGLT2i Initiators | 182 | 184989 | 0.98 (0.85, 1.14) |  |  |  |  |
| With Prior History of Suicidality |  |  |  |  |  |  |  |
| Suicide Ideation |  |  |  |  |  |  |  |
| Semaglutide Initiators | 848 | 4681 | 181 (170, 193) | 1.18 (1.10, 1.27) | P<.001* | 1.01 (0.93, 1.09) | P=0.82 |
| SGLT2i Initiators | 4452 | 30503 | 146 (142, 150) |  |  |  |  |
| Suicide Attempt/Completion |  |  |  |  |  |  |  |
| Semaglutide Initiators | 87 | 5942 | 14.6 (11.7, 18.0) | 1.34 (1.06, 1.69) | P=.014* | 1.08 (0.84, 1.39) | P=0.54 |
| SGLT2i Initiators | 405 | 37645 | 10.8 (9.7, 11.9) |  |  |  |  |

<sup>1</sup>HR = Hazard Ratio, CI = Confidence Interval

### Primary Outcomes

During a median follow-up of 2.2 years, 9077 and 696 total incident suicidal ideation events and suicide attempts/completion events occurred, respectively. For semaglutide and SGLT2i initiators with T2D, respectively, 1321 (11.5%) and 7756 (8.5%) reported suicidal ideation, and 109 (0.95%) and 587 (0.65%) had attempts/completions. The unadjusted incidence rate of suicidal ideation was 56.3 and 36.8 per 1,000 person-years for suicidal ideation and 4.30 and 2.64 per 1,000 person-years for attempts/completions among semaglutide and SGLT2i initiators with T2D.

### Intention-to-Treat Analyses

Unweighted Kaplan–Meier curves demonstrated early and persistent separation between groups, with the cumulative incidence of suicidal ideation and attempts/completions consistently higher among semaglutide initiators than SGLT2i initiators with T2D (Figure 1). In unweighted analyses, semaglutide initiators had an increased hazard of suicidal ideation and attempts/completions compared with SGLT2i initiators. Additionally, among patients without a history of suicidality, unweighted analyses demonstrated an increased risk of suicide ideation but not in attempts/completions in semaglutide initiators (Figure S3). Similarly, among patients with a history of suicidality, a significant increase in risk of suicide outcomes was observed in the unweighted analysis (Figure S4). However, after overlap weighting, none of these differences remained significant (Figure 1, Figure S3, and Figure S4).

**Figure 1.**
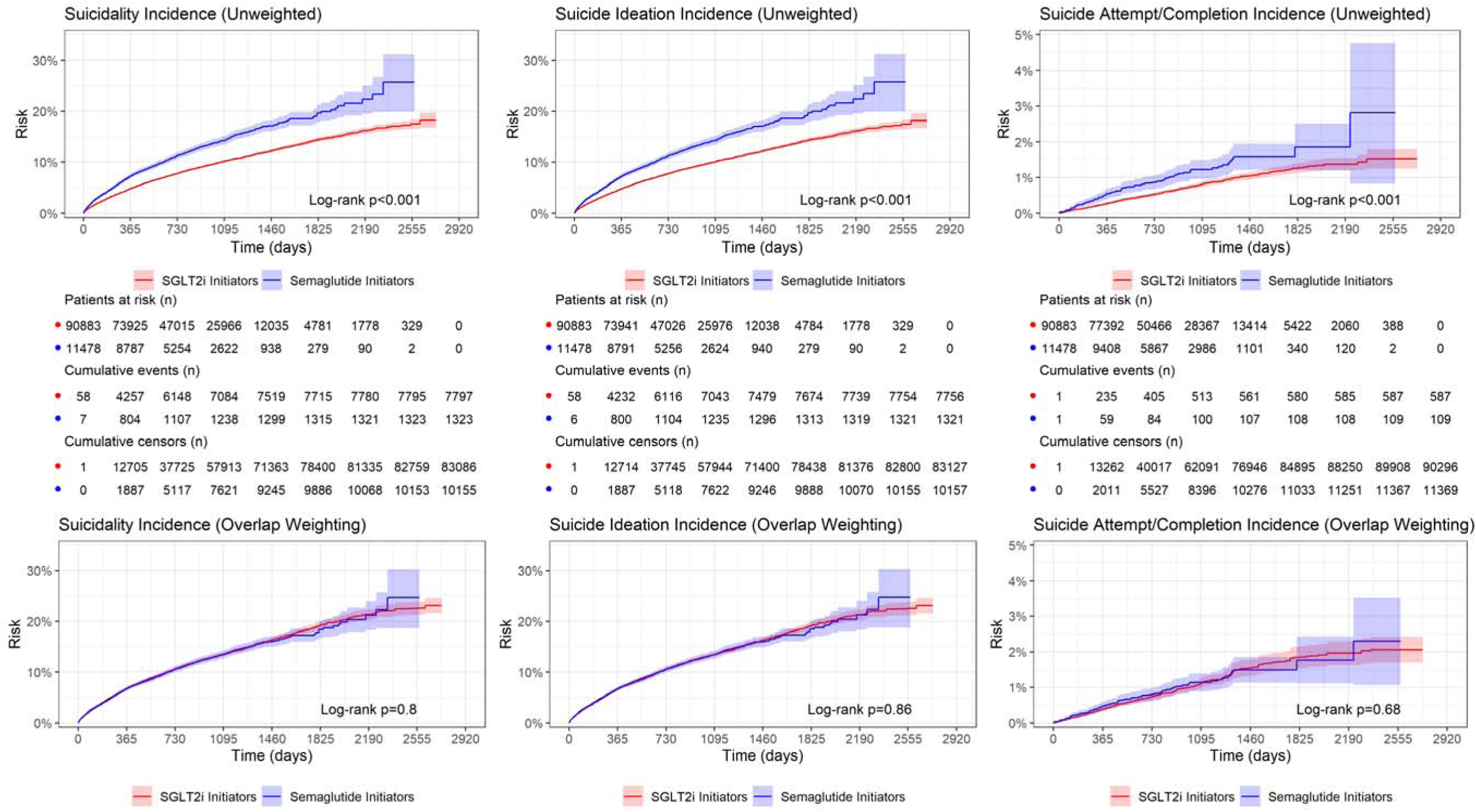
Risk Measure ITT Weighted Semaglutide vs SGLT2i for Suicide Outcomes. Kaplan-Meier cumulative incidence curves comparing semaglutide initiators (blue) with SGLT2i initiators (red) for three outcomes: composite suicidality (left), suicide ideation (center), and suicide attempt/completion (right). The top row presents unadjusted estimates; the bottom row presents estimates after overlap propensity score weighting to balance baseline characteristics between treatment groups. Shaded bands represent 95% confidence intervals. Log-rank p-values are displayed within each panel. In unadjusted analyses, semaglutide initiators had significantly higher cumulative incidence across all three outcomes (all log-rank P < .001), with separation between curves evident early and persisting throughout follow-up. After overlap weighting, the between-group differences were attenuated and no longer statistically significant for suicide ideation (log-rank P = 0.86), suicide attempt/completion (log-rank P = 0.68), or the composite outcome (log-rank P = 0.8), indicating that baseline confounders largely accounted for the observed crude differences. Numbers at risk, cumulative events, and cumulative censoring counts are displayed beneath each unweighted panel at evenly spaced time intervals. Follow-up extends to approximately 2,920 days (8 years). Abbreviations: SGLT2i, sodium-glucose cotransporter-2 inhibitor; ITT, intention-to-treat.

Findings were similar for the composite outcome for the overall cohort and when stratified by a history of suicidality at index (see Figure 1, Figure S3, and Figure S4).

### Per-Protocol Analyses with Clone Censored Weighting (CCW)

The results of the per-protocol analyses with the CCW method assess the predicted risk of primary suicide outcomes in three hypothetical scenarios in which all patients who initiated semaglutide adhered to each of the treatment arms. Using Arm A as reference, we observed a significant dose-response relationship between the length of adherence to semaglutide and suicidality outcomes (Figure 2). Compared to the short-term adherence (Arm A), the per-protocol effect of long-term adherence (Arm C) was associated with a 73% reduction in the hazard of suicide attempts/completions (HR, 0.27; 95% CI, 0.14–0.54; P<.001), and the intermediate-duration arm (Arm B) was associated with a non-significant reduction in risk. A duration-dependent effect was not observed for suicidal ideation.

**Figure 2.**
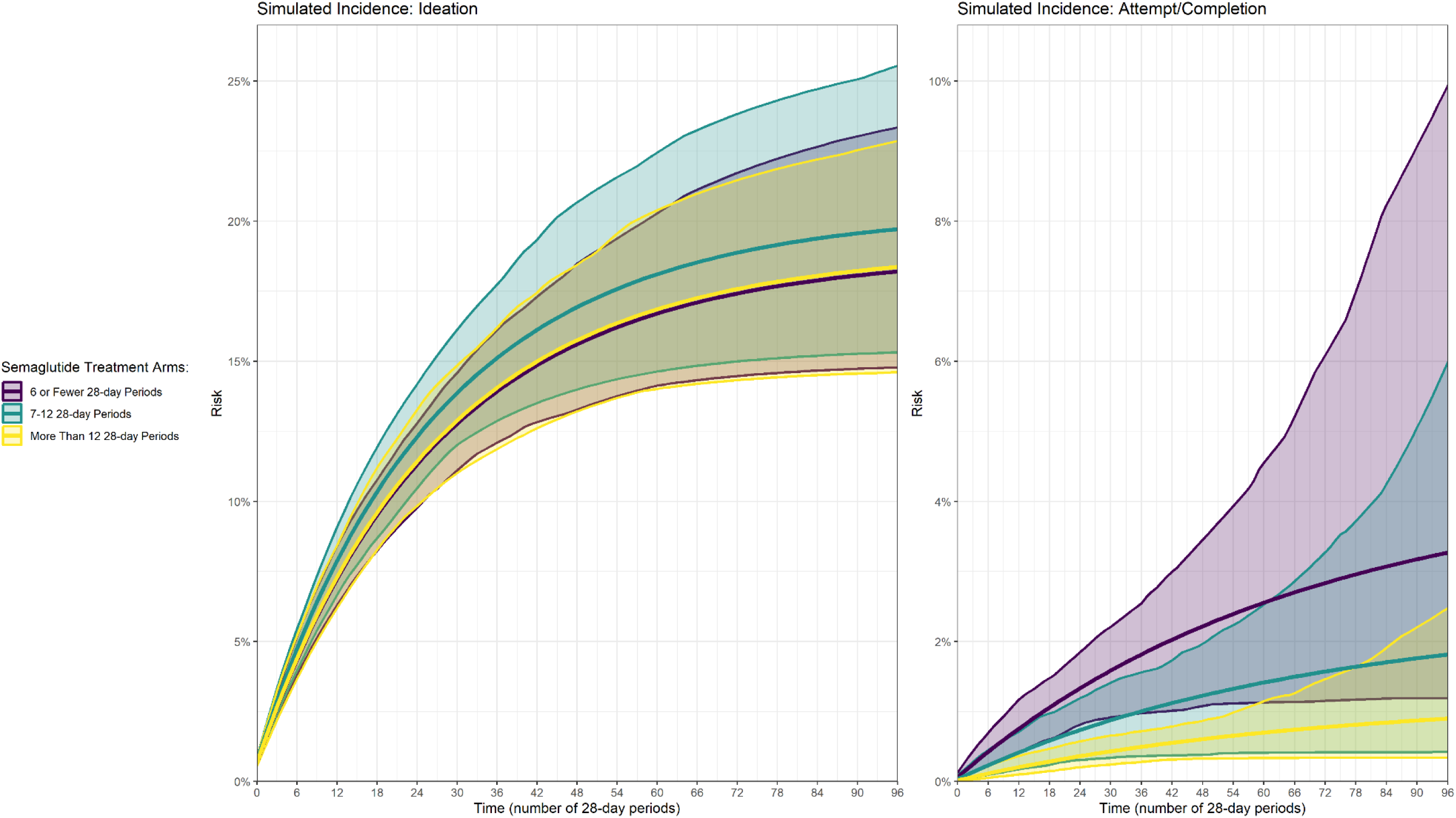
Clone Censor Weighting Simulation Results for Semaglutide Exposure Treatment Arms. Simulated average risk curves depicting the estimated cumulative incidence of suicide ideation (left) and suicide attempt/completion (right) under three hypothetical semaglutide treatment duration strategies among 11,478 semaglutide initiators: short-term adherence (Arm A: ≤6 twenty-eight-day periods [approximately ≤6 months]; purple), intermediate adherence (Arm B: 7–12 twenty-eight-day periods [approximately 6–12 months]; teal), and long-term adherence (Arm C: >12 twenty-eight-day periods [approximately >12 months]; yellow). Curves were generated using clone–censor–weight per-protocol methods, with inverse probability of adherence/censoring weights applied to adjust for selection bias introduced by artificial censoring. Shaded bands represent bootstrap-derived 95% confidence intervals based on 1,000 resampled iterations. The x-axis denotes time in twenty-eight-day periods (range 0–96, corresponding to approximately 7.4 years of follow-up). For suicide attempt/completion, sustained adherence beyond 12 months (Arm C) was associated with a significantly lower simulated risk compared with short-term adherence (Arm A) (HR, 0.27; 95% CI, 0.14–0.54; P < .001), with non-overlapping confidence bands observed over the first 57 periods. The intermediate-duration arm (Arm B) showed a non-significant reduction (HR, 0.56; 95% CI, 0.25–1.23; P = .15). For suicide ideation, no significant duration-dependent effect was observed, as reflected by the substantial overlap of confidence bands across all three treatment arms (Arm B: HR, 1.09; 95% CI, 0.87–1.37; P = .19; Arm C: HR, 1.01; 95% CI, 0.81–1.26; P = .89; reference: Arm A).

**Figure 3.**
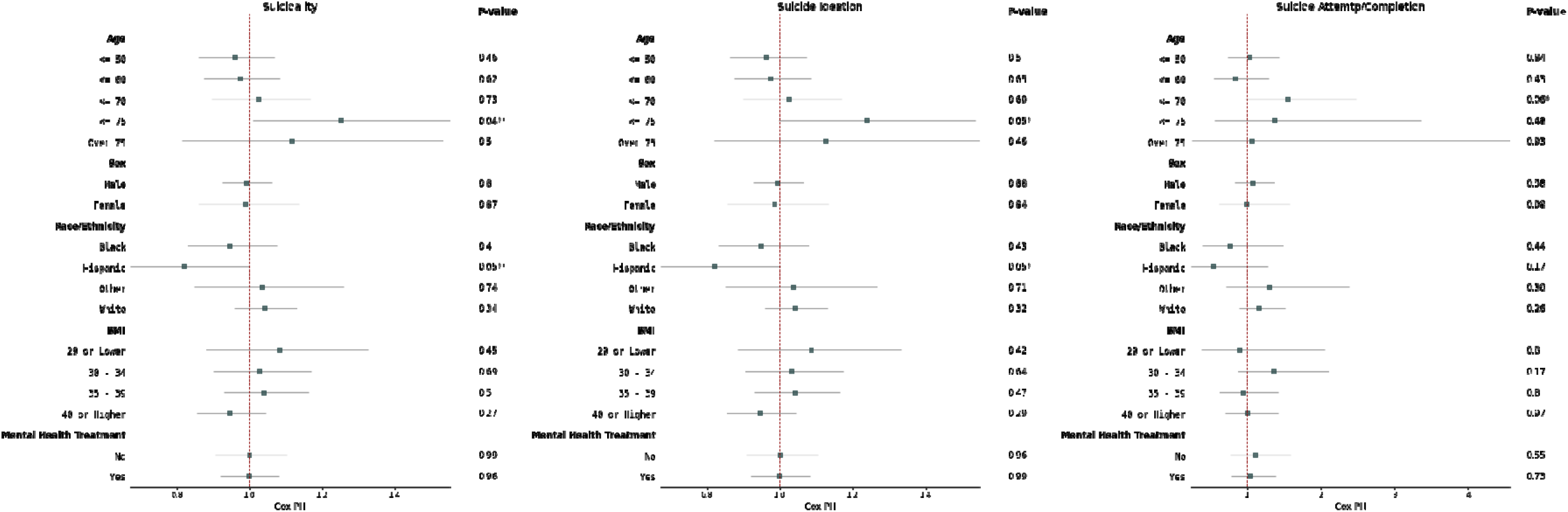
Forest Plot Showing the Effect of Semaglutide vs SGLT2 on Suicide Outcomes within Subgroups. Forest plot showing the comparative risks (x-axis) of combined suicidal ideation and attempt/completion outcomes (i.e. suicidality) (left), suicidal ideation outcomes (center), and suicide attempt/completion outcomes (right) in semaglutide versus SGLT2i initiators using overlap weighted stratified Cox proportional hazards ratio with 95% confidence intervals. The p-values from the Cox PH for each subgroup are shown in columns to the right of the bar plots. Increased risk of suicidality and ideation occurred in the patients aged 71–75 (p< .05) and decreased risk of suicidality occurred in patients with Hispanic ancestry (p< .05). Increased risk of suicide attempt/completion occurred in patients aged 61–70 (p<.05). No other significant effects were found for patients in the other subgroups by sex, BMI level, or concurrent mental health treatments.

These results are illustrated in the simulated average risk curves, i.e., the expected incidence of events if all patients were assigned and adherent to each treatment arm (Figure 2). The difference in average risk of suicide behavior between treatment arm A and treatment arm C is observed over the first 57 28-day periods; there is no overlap between the bootstrap resampled 95% CI during this interval. However, overlap is observed between the confidence bands for treatment arms A and B, and between treatment arms B and C. For suicidal ideation, the lack of an observed dose response across the treatment arms is illustrated by the near-complete overlap of the 95% CI around the simulated average event risk curves.

Patients ages 71–75 years and patients with Hispanic ancestry had a slightly increased risk of composite suicide outcome (i.e. suicidality (p=.04 and p=.05, respectively). A slight increased risk of suicide attempts/completion occurred in patients ages 61–70 years (p=.05). No subgroup reached the adjusted Bonferroni correction threshold α of 0.01 or 0.013, (5 comparisons across age groups, and 4 across race/ethnicity). No significant effects were observed based on sex, BMI, or concurrent mental health treatments.

## Discussion

In this nationwide target trial emulation of U.S. Veterans with T2D receiving metformin, semaglutide initiators were not observed to have an increased risk of suicidal ideation, suicide attempts/completion, or a composite suicidality outcome compared with SGLT2i initiators. Although unadjusted analyses suggested higher event rates among semaglutide initiators, overlap weighting in the ITT target trial attenuated the magnitude of this relationship, suggesting that baseline differences between treatment groups accounted for the unadjusted observations. In duration-of-treatment analyses using CCW methods to account for differential adherence and treatment discontinuation, longer-term semaglutide use was not associated with increased suicidality risk and was associated with lower risk of suicide attempts or death compared with shorter-term use. These findings suggest that suicidality risk does not accumulate with sustained exposure and may reflect adherence patterns or underlying differences in patient characteristics over time. Results were consistent across multiple subgroups and sensitivity analyses. Although residual confounding cannot be excluded, the concordance between initiation and adherence-adjusted analyses provides consistent evidence that semaglutide was not associated with elevated suicidality risk in this Veteran population after accounting for measured clinical differences.

Our findings align with recent meta-analyses of RCTs and observational cohort studies suggesting no aggregate link between semaglutide and suicidality.^3–5^ Additionally, our novel use of CCW methods provides critical granularity regarding the temporal nature of these risks. Our analysis revealed a significant dose-response relationship where longer periods of adherence were associated with a decreased risk of severe suicidality outcomes. Specifically, Veterans who remained on semaglutide for more than 12 months (Arm C) demonstrated a 73% lower hazard of suicide attempts or completions (HR: 0.27, p<0.001) compared to those with less than six months of exposure (Arm A). This duration-dependent protective effect was not observed for suicidal ideation, where risk remained stable across all adherence windows.

These findings may explain the inconsistencies currently found in the literature. Studies relying on pharmacovigilance reports or short follow-up periods^6^ likely capture patients primarily in the acute initiation phase (≤6 months), where we observed the highest relative risk per the per-protocol analysis of semaglutide exposure durations. Conversely, observational studies^8^ reporting protective effects may have captured more users with sustained adherence. Other explanations for inconsistencies may include differences in weighting strategies, comparator choice, use of new-user designs, length of follow-up, extent of confounder adjustment, and the rarity of suicidality.

Methodologically, this study highlights the importance of moving beyond ITT analogues in drug safety research. While our primary ITT analysis was reassuring, it essentially averaged risk across different adherence levels of treatment. Given that treatment duration is a post-baseline, time-varying exposure affected by both prior treatment and survival, the CCW method estimated the per-protocol effects of sustained semaglutide treatment strategies while appropriately accounting for time-varying confounding and informative censoring. In doing so, we were able to identify that the “neutral” risk in the overall cohort reflected an underlying composite of higher relative risk during early initiation and significantly lower risk during long-term maintenance.

These results are highly encouraging for clinicians treating Veterans, a population already at heightened risk for suicidality. The modelling results suggest that if a Veteran can successfully navigate the initial six months of semaglutide therapy and maintain adherence for 12 months, the long-term neuropsychiatric safety profile is excellent, with an ensuing potential reduction in the risk of suicide attempts. It is unclear if this protective effect remains past ∼57 months. Regardless, this study’s real-world evidence points to continued use of semaglutide as a longer term maintenance therapy for T2D in the Veteran population, even for those with a history of suicidality.

Future studies should determine if this protective effect persists beyond the ∼57 months found in our study. Additionally, future research should study such mechanisms and explore potential vulnerability factors, for example the impact of pharmacogenetic variants that impact the speed of metabolizing semaglutide. Future research may also consider examining changes in psychiatric symptoms in at-risk populations like those with mental health conditions, the effects of time-varying covariates like mental health treatment, and the effects of discontinuation of treatment.

This analysis leveraged the U.S.’s largest integrated health system, which offered extended follow-up (up to 7 years) and, by comparison, a relatively large number of incident suicidality events, including suicidal ideation, attempts and completions.

Methodologically, this study advances prior evidence in several ways. First, medication exposure was defined using outpatient pharmacy dispensing rather than prescription orders or administrative claims. Second, we incorporated detailed clinical covariates, including mental health history, history of suicidality, hemoglobin A1c, body mass index, blood pressure, and lipid levels. Third, our study statistical methods, including per-protocol analyses with CCW, have not been previously reported in the literature for this population and outcome.

## Limitations

Suicide attempts/completions remain rare events, leading to wider confidence intervals in stratified analyses. Additionally, underreporting of suicidality could affect overall incidence rates.

Our results in a predominantly male Veteran cohort with cardiometabolic burden may not be fully generalizable to the civilian population or to those using semaglutide solely for weight loss.

Despite our use of CCW to adjust for immortal time and baseline selection bias, potential for unmeasured confounders remains. For example, an association between a patient’s sensitivity to semaglutide (e.g., discontinuation due to intolerable side-effects) and predisposition to suicidality may exist via an unmeasured mechanism.

Patients may also stop adhering to the medication due to an increase in suicidality, although this explanation is less likely with our study’s censoring methods. (Supplemental Figure S2). Outcome events that occurred during the first period of non-adherence to a treatment arm counted towards the event rate of that treatment arm. For example, a patient with fewer than 14 days covered during period 3 (days 56-83 of follow-up) would have adherence to arm A and non-adherence to arms B and C. An outcome event occurring during period 3 would count toward the events rates of all 3 arms. If the event occurred on day 84 or later, the outcome event would only count towards the event rate of arm A because clones in arm B and arm C would have already been right censored. This effectively gives a 14-day grace period of non-adherence for events to count towards the event rates of all arms (towards the null) versus counting towards select arms (towards an effect).

Importantly, our study did not examine mechanisms for this differential effect based on adherence. Therefore, these findings must be interpreted with caution. Discontinuing semaglutide may possibly be itself a risk factor for suicidality and as such, patients discontinuing treatment with less than 6 months of adherence would have more post-discontinuance at-risk time compared to patients end of follow-up in the long-term adherence arm. Further research is needed to determine directionality.

## Conclusions

Among Veterans with T2D, semaglutide initiators were not observed to have higher risks of suicidal ideation, suicide attempts/completions, or a composite outcome compared with initiation SGLT2i initiators. This held true even in populations at higher risk for suicidality. Crucially, semaglutide initiators who sustained adherence to semaglutide for more than one year had lower risks of suicide attempts and completions. These findings provide evidence that longer term use of semaglutide is safe and potentially protective against severe suicidality outcomes in high-risk populations.

## Data Availability

Patient-level data are accessible to all VA researchers with appropriate IRB approvals.

## Acknowledgements

This work was supported by using resources and facilities of the Department of Veterans Affairs (VA) Informatics and Computing Infrastructure (VINCI), including manuscript preparation by Kathryn Pridgen, which is funded under the research priority to Put VA Data to Work for Veterans (VA ORD 24-D4V-02).

## Disclaimer

The views expressed in this article are those of the authors and do not necessarily reflect the position or policy of the Department of Veterans Affairs or the United States government.

