## Supplemental Material for "Semaglutide Initiation and Treatment Duration On Suicidality Risk in US Veterans With Type 2 Diabetes"

Supplemental Methods

CCW methods were implemented as follows:

- **Step 1: Cloning.** At time zero, we created three identical copies ("clones") of each semaglutide initiator. One clone was assigned to each of the three treatment duration strategies (arms A, B, and C): A) initiation and continuous exposure to semaglutide for ≤6 28-day periods, B) initiation and continuous exposure to semaglutide for 7–12 28-day periods, and C) initiation and continuous exposure to semaglutide for >12 28-day periods.
- **Step 2: Artificial Censoring.** We monitored each clone’s medication possession over time. A clone was right censored for the period during which their real-world data deviated from their assigned strategy. Participants in arm A were censored if they had continuous exposure for 7 28-day periods, or if they were re-exposed after stopping during the first 6 28-day periods, whichever occurred first. Participants in arm B were censored if continuous exposure was not met during any of the first 6 28-day periods, if continuous exposure continued for 13 28-day periods, or if they were re-exposed after stopping during periods 7–12, whichever occurred first. Participants in arm C were censored if continuous exposure was not met during any of the first 12 28-day periods or if they were re-exposed after stopping exposure after period 13.
- **Step 3:** **Weighting.** To account for potential selection bias introduced by informative censoring (patients who discontinue may differ from those who remain adherent), we calculated unstabilized inverse probability of adherence/censoring weights (IPAW/IPCW) for each treatment arm and period based on the predicted probability of an individual being adherent/censored to the treatment protocol conditional on baseline covariates. Values were truncated to the 99^th^ percentile.

Supplemental Tables and Figures

Table S1. Target Trial Specification and Emulation

| Approaches | Target Trial | Target Trial Emulation |
| --- | --- | --- |
| Eligibility criteria | Target trial among individuals who meet the following inclusion criteria:   - Age at least 18 at index date - T2D - Active Metformin Rx in past 90 days - At least 1 of any the following within 1 year prior to index date: - Outpatient workload visit - Outpatient prescription - Laboratory test for blood glucose or HbA1c   Exclude patients who meet the following exclusion criteria:   - Prescription for Semaglutide or any other GLP-1RA ever - Prescription for SGLT2 ever - Diagnosis code(s) for any of the following within 1 year of index date: - Type 1 Diabetes or Gestational Diabetes | Same as for target trial |
| Treatment strategies | Intervention arm: Treatment initiation of  Semaglutide  Comparator Control arm: Treatment initiation of SGLT2 | Same as for target trial |
| Treatment assignments | Individuals are randomly assigned to a strategy at baseline. Individuals and their treating physicians will be aware of the assigned treatment strategy. | We defined the date of medication initiation to be the first date of a prescription.  Inverse propensity treatment weighted derived from propensity score calculated based on baseline covariates to balance the characteristic between treatment groups. |
| Outcomes | **Suicide (binary):**  Combined outcome: suicidal ideation, attempts, and/or completion  Suicidal Ideation only: suicidal ideation  Suicidal Behaviors: attempt and/or completion | Same as for target trial. |
| Follow-up | At least 6 months of follow-up | Same as for target trial. |
| Causal contrasts | Intention-to-treat  Per-protocol for adherence to Semaglutide |  |
| Statistical analysis | For incident outcomes, Cox proportional hazards regression model to estimate hazard ratios and KaplanMeier plot to show the cumulative incidence over time with overlap weighting  For treatment adherence, per-protocol analysis    Subgroup analyses by 1) history of mental illness (yes or no); 2) mental health treatment (yes or no) | Same as for the target trial  Additional sensitivity  analyses: 1) using a 1:1  propensity score matched  Cox model 2) using a per-protocol analysis |

Table S2. Definitions for Covariates

| Variable | Definition |
| --- | --- |
| ICD-10 Codes | |
| Depression | F32-F33 |
| Mood Disorders, including bipolar disorder | F30-F39 |
| Anxiety | F40-48 |
| Behavioral Disorders | F50-59 |
| Personality Disorders | F60-F69 |
| Psychotic Disorders | F20-F29 |
| PTSD | F43.1 (F43.10-F43.12) |
| Substance use disorders | F10-19 (Wang et al excluded 13, 16,18) |
| Chronic pain | G89.2 |
| Sleep disorders | G47; sleep disorder not due to substance or physiological condition (F51) |
| TBI | S06 |
| Cancer | C00-D49 |
| Bariatric Surgery | Z98.84 |
| History of psychological trauma | Z91.4 |
| Family History of Mental disorders | Z81 |
| Adverse socioeconomic and psychosocial Problems | Persons with potential health hazards related to socioeconomic and psychosocial circumstances (ICD-10 code: Z55-65): including Problems related to education (Z55), employment Z56), housing (Z59), social environment (Z60), upbringing (Z62), family circumstances (Z63), psychosocial circumstance s(Z64,Z65) |
| Lifestyle Problems | Z72 |
| Medications | |
| Diabetes Medications | Insulins and analogues (ATC code: A10A), Metformin (RxNorm code: 6809), Sulfonylureas (ATC code: 6809), Alpha glucosidase inhibitors (ATC code: A10BH), Thiazolidinedione (ATC code: A10BG), DPP-4 Inhibitor (ATC code: A10BH), SGLT2 inhibitor (ATC code: A10BK) |
| Mental Health Medications | Antidepressants (ATC code: N06A), Antipsychotics (ATC code: N05A), Antiepileptics (ATC code: N03), Benzodiazepine derivatives/hypnotics (VA code: CN302), Esketamine (RxNorm code: 2119365), ketamine (RxNorm code: 6130), Lithium (RxNorm code: 6448) |
| Treatment (CPT Codes) | |
| Psychotherapy | Individual (90832, 90834, 90837), group therapy (90853), couples/family therapy (90847, 90849), crises interventions (S9484, H2011, 90839, 90840) |

Table S3. Absolute Incidence Rates and Hazard Ratios of Suicide Ideation in Unweighted and Overlap Weighted Sub-Cohorts

|  |  |  |  |  |  | **Unweighted** |  | **Overlap Weighted** |  |
| --- | --- | --- | --- | --- | --- | --- | --- | --- | --- |
|  |  | **N** | **Events** | **At-Risk Time (years)** | **Incidence Rate (95% CI) per 1000 Person-Years** | **HR***^1^* **(95% CI)** | **p-value** | **HR***^1^* **(95% CI)** | **p-value** |
| Age Quintile |  |  |  |  |  |  |  |  |  |
| ≤50 | Semaglutide | 2673 | 450 | 5114 | 88.0 (80.4, 96.1) | 1.15 (1.04, 1.28) | P=0.0072** | 0.96 (0.86, 1.07) | P=0.50 |
|  | SGLT2i | 12099 | 1938 | 26631 | 72.8 (69.7, 76.0) |  |  |  |  |
| 51–60 | Semaglutide | 3525 | 439 | 7042 | 62.3 (56.8, 68.2) | 1.15 (1.03, 1.27) | P=0.0090** | 0.98 (0.88, 1.09) | P=0.65 |
|  | SGLT2i | 19700 | 2376 | 45633 | 52.1 (50.0, 54.1) |  |  |  |  |
| 61–70 | Semaglutide | 3005 | 286 | 6382 | 44.8 (39.9, 50.2) | 1.32 (1.17, 1.50) | P<.001*** | 1.03 (0.90, 1.17) | P=0.69 |
|  | SGLT2i | 24919 | 1956 | 60154 | 32.5 (31.1, 34.0) |  |  |  |  |
| 71–75 | Semaglutide | 1266 | 102 | 2944 | 34.7 (28.3, 41.9) | 1.60 (1.31, 1.97) | P<.001*** | 1.24 (1.00, 1.54) | P=0.051 |
|  | SGLT2i | 16878 | 917 | 43611 | 21.0 (19.7, 22.4) |  |  |  |  |
| >75 | Semaglutide | 1009 | 44 | 1973 | 22.3 (16.2, 29.8) | 1.35 (0.99, 1.83) | P=0.058 | 1.13 (0.82, 1.55) | P=0.46 |
|  | SGLT2i | 17287 | 569 | 34449 | 16.5 (15.2, 17.9) |  |  |  |  |
| Birth sex |  |  |  |  |  |  |  |  |  |
| Male | Semaglutide | 9500 | 1011 | 19527 | 51.8 (48.7, 55.0) | 1.41 (1.32, 1.51) | P<.001*** | 0.99 (0.93, 1.07) | P=0.88 |
|  | SGLT2i | 85134 | 6995 | 197850 | 35.4 (34.5, 36.2) |  |  |  |  |
| Female | Semaglutide | 1978 | 310 | 3928 | 78.9 (70.7, 87.8) | 1.28 (1.12, 1.46) | P<.001*** | 0.99 (0.86, 1.13) | P=0.84 |
|  | SGLT2i | 5749 | 761 | 12627 | 60.3 (56.2, 64.6) |  |  |  |  |
| Race/ethnicity |  |  |  |  |  |  |  |  |  |
| Black | Semaglutide | 2480 | 316 | 4769 | 66.3 (59.4, 73.7) | 1.28 (1.14, 1.44) | P<.001*** | 0.95 (0.83, 1.08) | P=0.43 |
|  | SGLT2i | 16218 | 1766 | 35814 | 49.3 (47.1, 51.6) |  |  |  |  |
| Hispanic | Semaglutide | 948 | 129 | 1791 | 72.0 (60.5, 85.0) | 1.17 (0.97, 1.40) | P=0.10 | 0.82 (0.67, 1.00) | P=0.052 |
|  | SGLT2i | 6951 | 917 | 15415 | 59.5 (55.8, 63.3) |  |  |  |  |
| Other | Semaglutide | 1058 | 127 | 2152 | 59.0 (49.4, 69.8) | 1.38 (1.15, 1.67) | P<.001*** | 1.04 (0.85, 1.27) | P=0.71 |
|  | SGLT2i | 8896 | 817 | 19705 | 41.5 (38.7, 44.3) |  |  |  |  |
| Non-Hispanic White | Semaglutide | 6992 | 749 | 14743 | 50.8 (47.3, 54.5) | 1.61 (1.49, 1.74) | P<.001*** | 1.04 (0.96, 1.13) | P=0.32 |
|  | SGLT2i | 58818 | 4256 | 139543 | 30.5 (29.6, 31.4) |  |  |  |  |
| Body mass index |  |  |  |  |  |  |  |  |  |
| <30 | Semaglutide | 1065 | 100 | 2435 | 41.1 (33.5, 49.7) | 1.38 (1.13, 1.69) | P=0.0017** | 1.09 (0.89, 1.33) | P=0.42 |
|  | SGLT2i | 27579 | 1854 | 62251 | 29.8 (28.5, 31.1) |  |  |  |  |
| 30–34.9 | Semaglutide | 2632 | 261 | 5561 | 46.9 (41.5, 52.8) | 1.33 (1.17, 1.51) | P<.001*** | 1.03 (0.90, 1.18) | P=0.64 |
|  | SGLT2i | 30154 | 2422 | 70711 | 34.3 (32.9, 35.6) |  |  |  |  |
| 35–39.9 | Semaglutide | 3320 | 397 | 6665 | 59.6 (54.0, 65.5) | 1.40 (1.25, 1.56) | P<.001*** | 1.04 (0.93, 1.17) | P=0.47 |
|  | SGLT2i | 19690 | 1883 | 46230 | 40.7 (38.9, 42.6) |  |  |  |  |
| ≥40 | Semaglutide | 4461 | 563 | 8795 | 64.0 (59.0, 69.3) | 1.19 (1.08, 1.31) | P<.001*** | 0.95 (0.86, 1.05) | P=0.29 |
|  | SGLT2i | 13460 | 1597 | 31286 | 51.0 (48.6, 53.5) |  |  |  |  |
| Concurrent mental health treatments |  |  |  |  |  |  |  |  |  |
| No | Semaglutide | 7771 | 504 | 16745 | 30.1 (27.6, 32.8) | 1.34 (1.22, 1.47) | P<.001*** | 1.00 (0.91, 1.11) | P=0.96 |
|  | SGLT2i | 72293 | 3799 | 172093 | 22.1 (21.4, 22.8) |  |  |  |  |
| Yes | Semaglutide | 3707 | 817 | 6710 | 121.8 (114.0, 129.8) | 1.12 (1.04, 1.21) | P=0.0035** | 1.00 (0.92, 1.08) | P=0.99 |
|  | SGLT2i | 18590 | 3957 | 38384 | 103.1 (100.1, 106.2) |  |  |  |  |

Table S4. Absolute Incidence Rates and Hazard Ratios of Suicide Attempt/completion in Unweighted and Overlap Weighted Sub-Cohorts

|  |  |  |  |  |  | **Unweighted** |  | **Overlap Weighted** |  |
| --- | --- | --- | --- | --- | --- | --- | --- | --- | --- |
|  |  | **N** | **Events** | **At-Risk Time (years)** | **Incidence Rate (95% CI) per 1000 Person-Years** | **HR***^1^* **(95% CI)** | **p-value** | **HR***^1^* **(95% CI)** | **p-value** |
| Age quintile |  |  |  |  |  |  |  |  |  |
| ≤50 | Semaglutide | 2673 | 49 | 5728 | 8.6 (6.3, 11.3) | 1.24 (0.91, 1.70) | P=0.18 | 1.03 (0.74, 1.45) | P=0.84 |
|  | SGLT2i | 12099 | 201 | 29755 | 6.8 (5.9, 7.8) |  |  |  |  |
| 51–60 | Semaglutide | 3525 | 28 | 7704 | 3.6 (2.4, 5.2) | 0.96 (0.65, 1.44) | P=0.86 | 0.84 (0.54, 1.30) | P=0.43 |
|  | SGLT2i | 19700 | 183 | 49433 | 3.7 (3.2, 4.3) |  |  |  |  |
| 61–70 | Semaglutide | 3005 | 24 | 6790 | 3.5 (2.3, 5.3) | 1.80 (1.16, 2.78) | P=0.0088** | 1.55 (0.97, 2.48) | P=0.064 |
|  | SGLT2i | 24919 | 123 | 63194 | 1.9 (1.6, 2.3) |  |  |  |  |
| 71–75 | Semaglutide | 1266 | 6 | 3105 | 1.9 (0.7, 4.2) | 2.00 (0.85, 4.69) | P=0.11 | 1.38 (0.56, 3.37) | P=0.48 |
|  | SGLT2i | 16878 | 43 | 45123 | 1.0 (0.7, 1.3) |  |  |  |  |
| >75 | Semaglutide | 1009 | 2 | 2030 | 1.0 (0.1, 3.6) | 0.94 (0.23, 3.90) | P=0.93 | 1.06 (0.25, 4.56) | P=0.93 |
|  | SGLT2i | 17287 | 37 | 35128 | 1.1 (0.7, 1.5) |  |  |  |  |
| Birth sex |  |  |  |  |  |  |  |  |  |
| Male | Semaglutide | 9500 | 78 | 20952 | 3.7 (2.9, 4.6) | 1.46 (1.15, 1.85) | P=0.0018** | 1.07 (0.84, 1.38) | P=0.58 |
|  | SGLT2i | 85134 | 526 | 208827 | 2.5 (2.3, 2.7) |  |  |  |  |
| Female | Semaglutide | 1978 | 31 | 4407 | 7.0 (4.8, 10.0) | 1.58 (1.02, 2.43) | P=0.039* | 0.99 (0.62, 1.59) | P=0.98 |
|  | SGLT2i | 5749 | 61 | 13807 | 4.4 (3.4, 5.7) |  |  |  |  |
| Race/ethnicity |  |  |  |  |  |  |  |  |  |
| Black | Semaglutide | 2480 | 12 | 5261 | 2.3 (1.2, 4.0) | 0.97 (0.53, 1.78) | P=0.93 | 0.77 (0.40, 1.50) | P=0.44 |
|  | SGLT2i | 16218 | 89 | 38611 | 2.3 (1.9, 2.8) |  |  |  |  |
| Hispanic | Semaglutide | 948 | 7 | 1967 | 3.6 (1.4, 7.3) | 0.82 (0.38, 1.77) | P=0.61 | 0.55 (0.24, 1.29) | P=0.17 |
|  | SGLT2i | 6951 | 72 | 16887 | 4.3 (3.3, 5.4) |  |  |  |  |
| Other | Semaglutide | 1058 | 14 | 2315 | 6.0 (3.3, 10.1) | 1.64 (0.93, 2.91) | P=0.088 | 1.31 (0.72, 2.39) | P=0.38 |
|  | SGLT2i | 8896 | 76 | 20905 | 3.6 (2.9, 4.5) |  |  |  |  |
| Non-Hispanic White | Semaglutide | 6992 | 76 | 15816 | 4.8 (3.8, 6.0) | 1.99 (1.55, 2.55) | P<.001*** | 1.17 (0.89, 1.53) | P=0.26 |
|  | SGLT2i | 58818 | 350 | 146231 | 2.4 (2.1, 2.7) |  |  |  |  |
| Body mass index |  |  |  |  |  |  |  |  |  |
| <30 | Semaglutide | 1065 | 6 | 2579 | 2.3 (0.9, 5.1) | 1.22 (0.54, 2.76) | P=0.64 | 0.90 (0.39, 2.06) | P=0.80 |
|  | SGLT2i | 27579 | 124 | 65107 | 1.9 (1.6, 2.3) |  |  |  |  |
| 30–34.9 | Semaglutide | 2632 | 24 | 5928 | 4.0 (2.6, 6.0) | 1.73 (1.13, 2.65) | P=0.012* | 1.36 (0.88, 2.11) | P=0.17 |
|  | SGLT2i | 30154 | 173 | 74514 | 2.3 (2.0, 2.7) |  |  |  |  |
| 35–39.9 | Semaglutide | 3320 | 30 | 7245 | 4.1 (2.8, 5.9) | 1.19 (0.80, 1.75) | P=0.39 | 0.95 (0.63, 1.42) | P=0.80 |
|  | SGLT2i | 19690 | 169 | 49149 | 3.4 (2.9, 4.0) |  |  |  |  |
| ≥40 | Semaglutide | 4461 | 49 | 9605 | 5.1 (3.8, 6.7) | 1.41 (1.01, 1.97) | P=0.043* | 1.01 (0.71, 1.43) | P=0.97 |
|  | SGLT2i | 13460 | 121 | 33864 | 3.6 (3.0, 4.3) |  |  |  |  |
| Concurrent mental health treatments |  |  |  |  |  |  |  |  |  |
| No | Semaglutide | 7771 | 39 | 17442 | 2.2 (1.6, 3.1) | 1.47 (1.05, 2.06) | P=0.025* | 1.12 (0.78, 1.59) | P=0.55 |
|  | SGLT2i | 72293 | 271 | 177797 | 1.5 (1.3, 1.7) |  |  |  |  |
| Yes | Semaglutide | 3707 | 70 | 7916 | 8.8 (6.9, 11.2) | 1.22 (0.94, 1.58) | P=0.13 | 1.05 (0.79, 1.39) | P=0.73 |
|  | SGLT2i | 18590 | 316 | 44837 | 7.0 (6.3, 7.9) |  |  |  |  |

Figure S1. CONSORT Cohort Selection

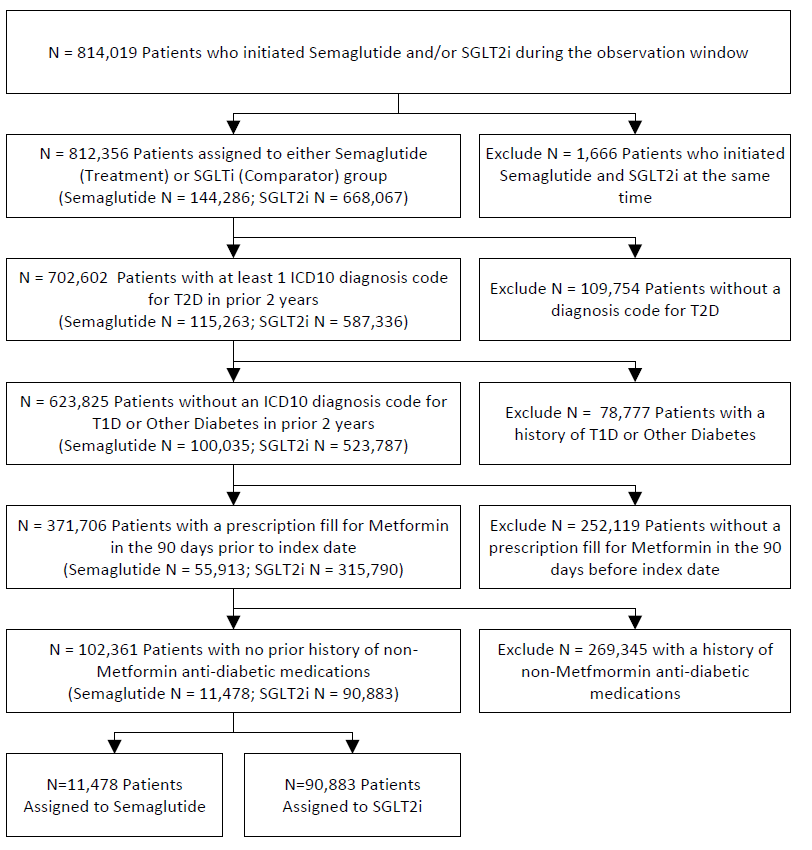

Figure S2. Clone Censor Weighting Schematic (Diagram.xlsx)

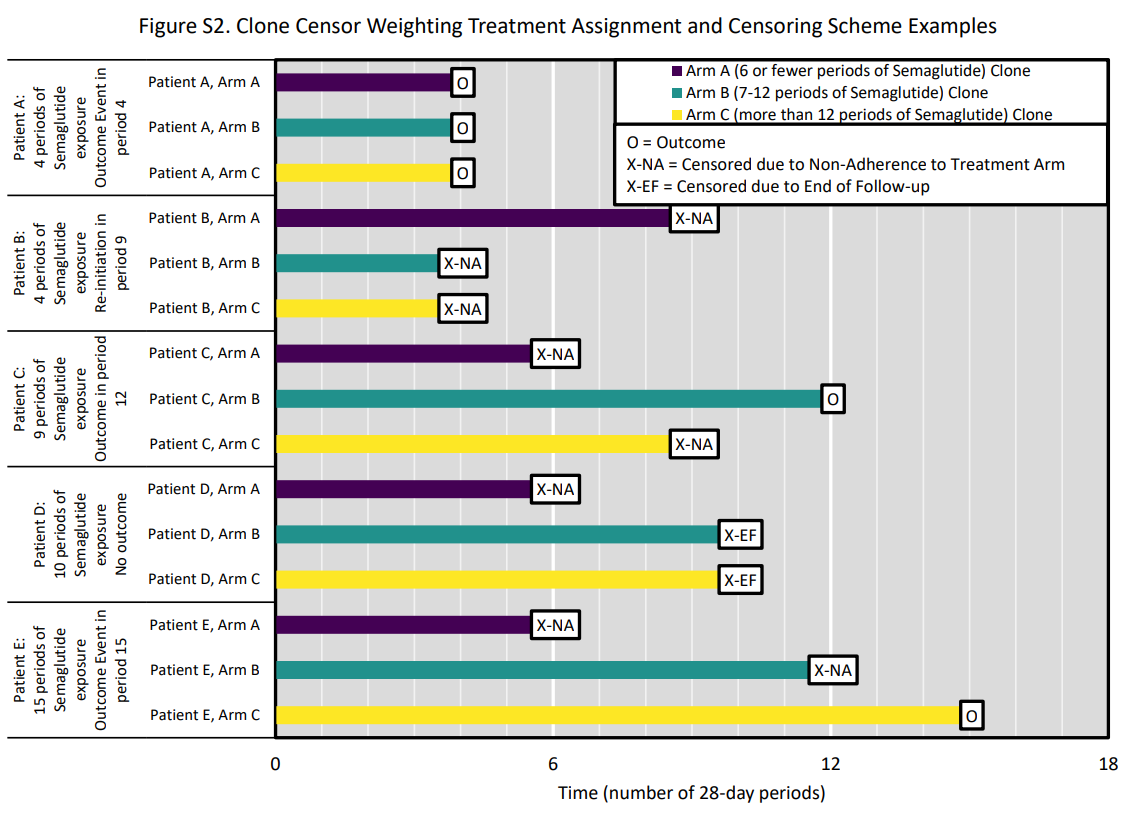

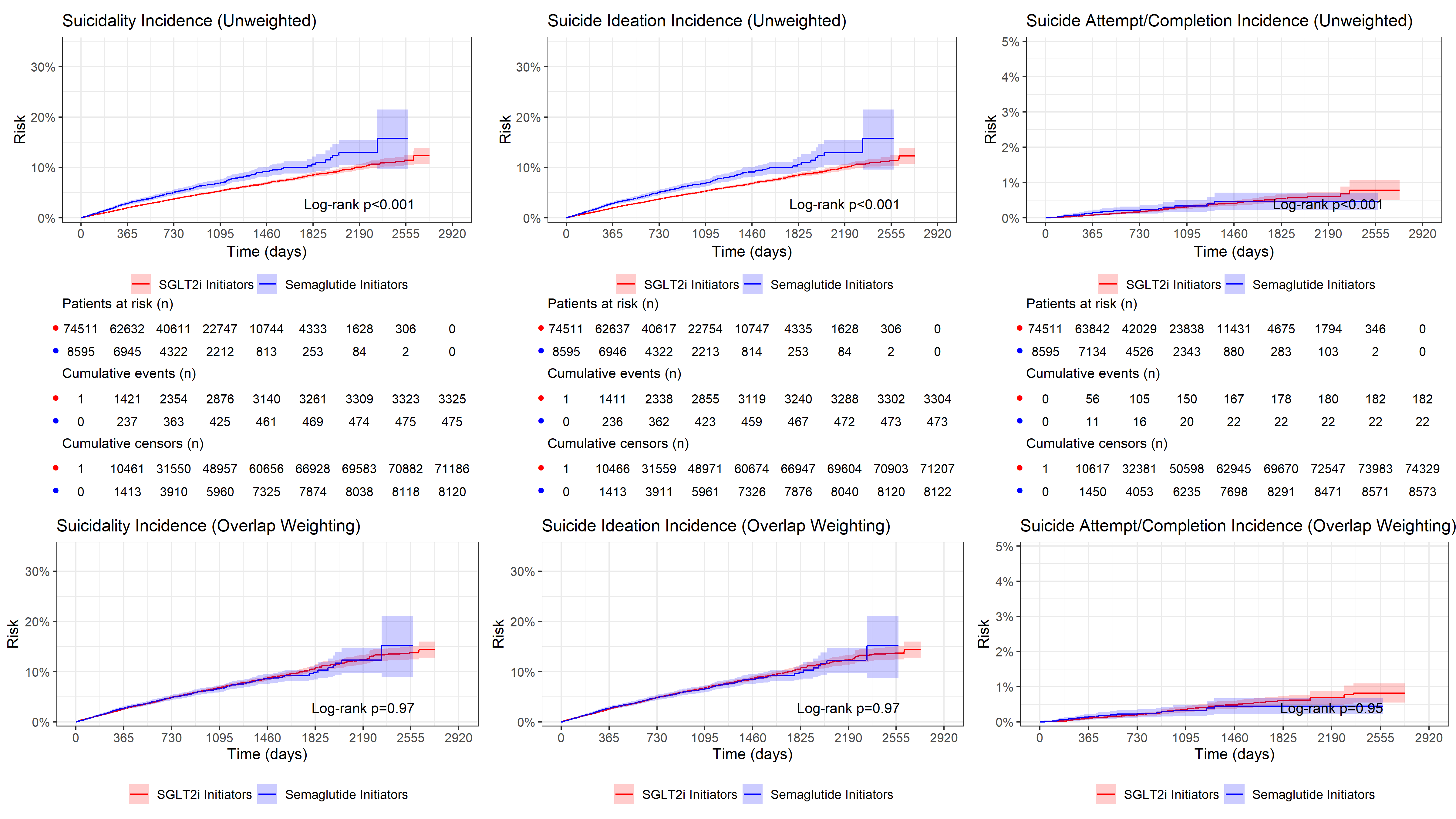
Figure S3. Risk Measure ITT Weighted Semaglutide vs SGLT2i for Suicide Outcomes Among Patients Without Prior History of Suicidality

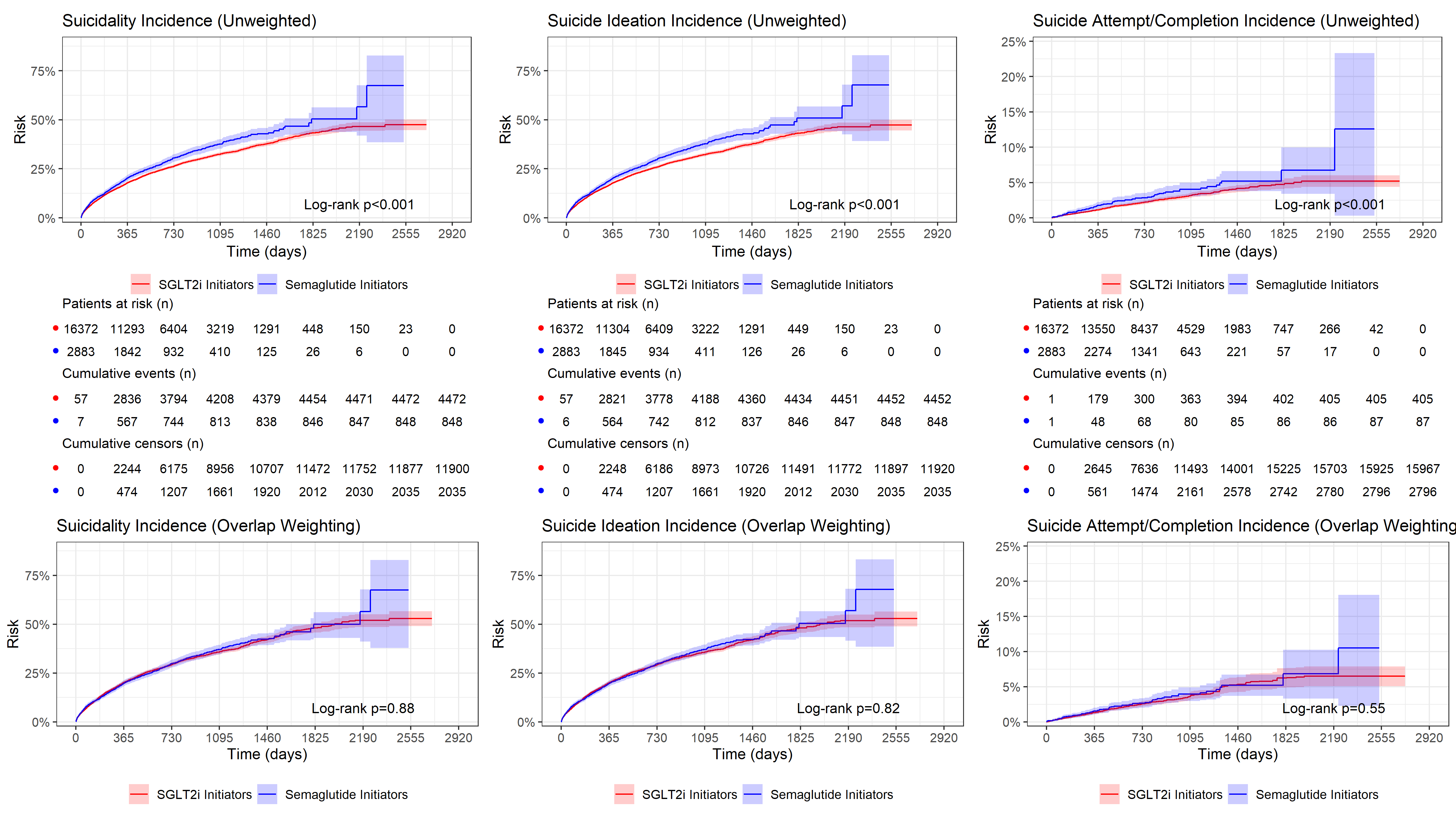
Figure S4. Risk Measure ITT Weighted Semaglutide vs SGLT2i for Suicide Outcomes Among Patients With Prior History of Suicidality

Figure Legend: Kaplan-Meier cumulative incidence curves comparing semaglutide initiators (blue) with SGLT2i initiators (red) among the subgroup of veterans with a prior history of suicidality (n = 2,883 semaglutide; n = 16,372 SGLT2i) for three outcomes: composite suicidality (left), suicide ideation (center), and suicide attempt/completion (right). The top row presents unadjusted estimates; the bottom row presents estimates after overlap propensity score weighting. Shaded bands represent 95% confidence intervals. Log-rank p-values are displayed within each panel. Numbers at risk, cumulative events, and cumulative censoring counts are shown beneath each unweighted panel at evenly spaced time intervals. In this high-risk subgroup, cumulative incidence was substantially higher than in the overall cohort, with unadjusted suicide ideation exceeding 50% and suicide attempt/completion approaching 15% among semaglutide initiators over the follow-up period. Unadjusted analyses showed significantly higher cumulative incidence among semaglutide initiators for all three outcomes (all log-rank P < .001), with early and persistent separation between curves. After overlap weighting, between-group differences were fully attenuated and non-significant for composite suicidality (log-rank P = 0.88), suicide ideation (log-rank P = 0.82), and suicide attempt/completion (log-rank P = 0.55), consistent with the primary analysis and indicating that measured baseline confounders accounted for the crude associations even among veterans at elevated baseline risk. Follow-up extends to approximately 2,920 days (8 years). Abbreviations: SGLT2i, sodium-glucose cotransporter-2 inhibitor; ITT, intention-to-treat.

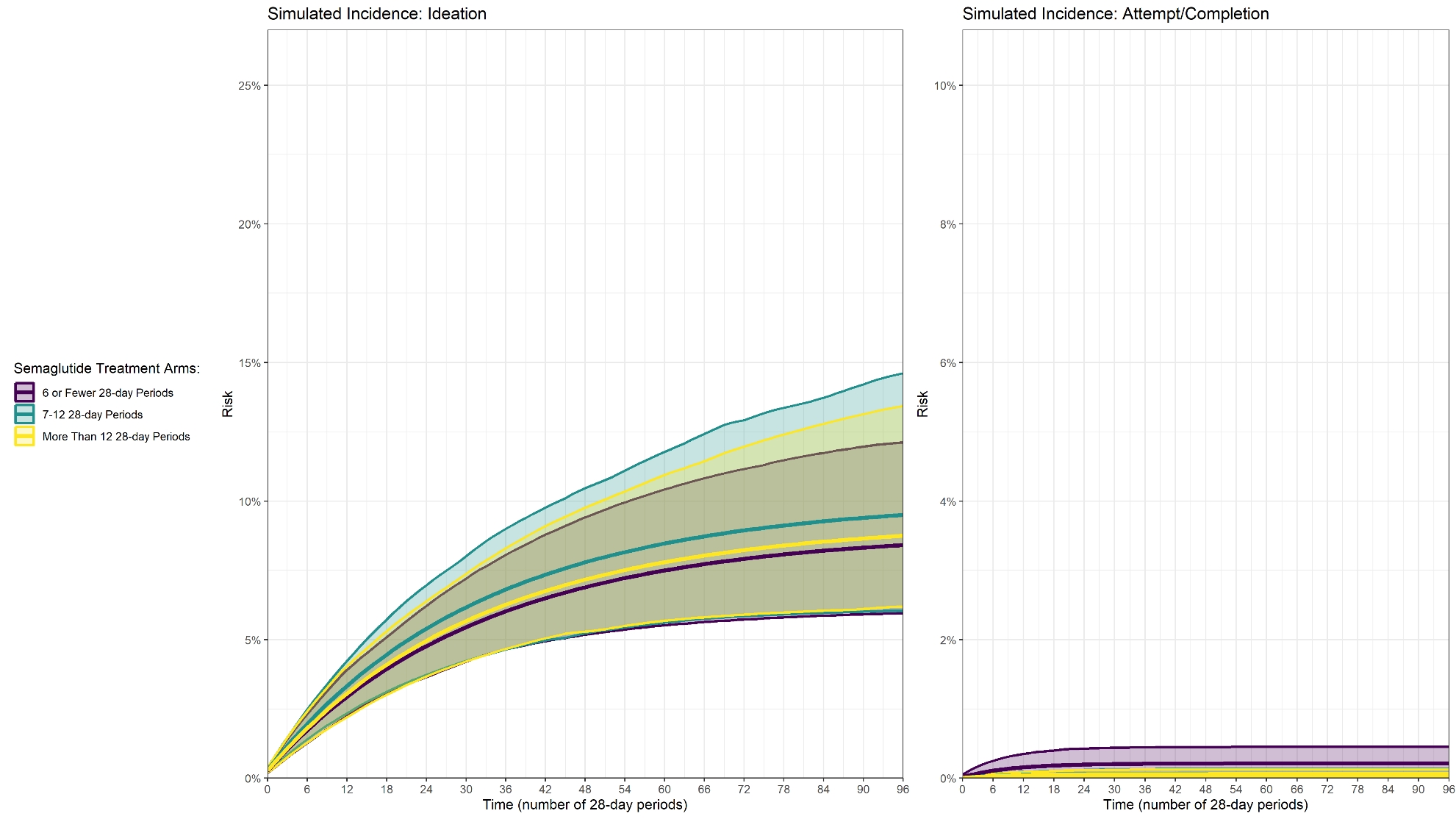
Figure S5. Risk Measure Clone Censor Weighting Simulation Results Among Patients Without a Prior History of Suicidality

Figure legend: Simulated average risk curves depicting the estimated cumulative incidence of suicide ideation (left) and suicide attempt/completion (right) under three hypothetical semaglutide treatment duration strategies among the subgroup of semaglutide initiators without a prior history of suicidality: short-term adherence (Arm A: ≤6 twenty-eight-day periods [approximately ≤6 months]; purple), intermediate adherence (Arm B: 7–12 twenty-eight-day periods [approximately 6–12 months]; teal), and long-term adherence (Arm C: >12 twenty-eight-day periods [approximately >12 months]; yellow). Curves were generated using clone–censor–weight per-protocol methods, with inverse probability of adherence/censoring weights applied to adjust for selection bias introduced by artificial censoring. Shaded bands represent bootstrap-derived 95% confidence intervals based on 1,000 resampled iterations. The x-axis denotes time in twenty-eight-day periods (range 0–96, corresponding to approximately 7.4 years of follow-up). In this subgroup without prior suicidality, the overall cumulative incidence was markedly lower than in the full cohort, with suicide ideation reaching approximately 6–11% and suicide attempt/completion remaining below 1% across all treatment arms by the end of follow-up. For suicide ideation, modest separation between the three treatment duration curves was observed, with the intermediate-duration arm (Arm B, teal) showing the highest simulated risk, though confidence bands overlapped substantially across all arms. For suicide attempt/completion, cumulative incidence remained very low across all three treatment arms, with all curves clustered near 0% and broadly overlapping confidence intervals, consistent with the rarity of these events among veterans without a prior history of suicidality. The x-axis denotes time in twenty-eight-day periods (range 0–96). Abbreviations: Arm A, ≤6 months semaglutide adherence; Arm B, 6–12 months; Arm C, >12 months.

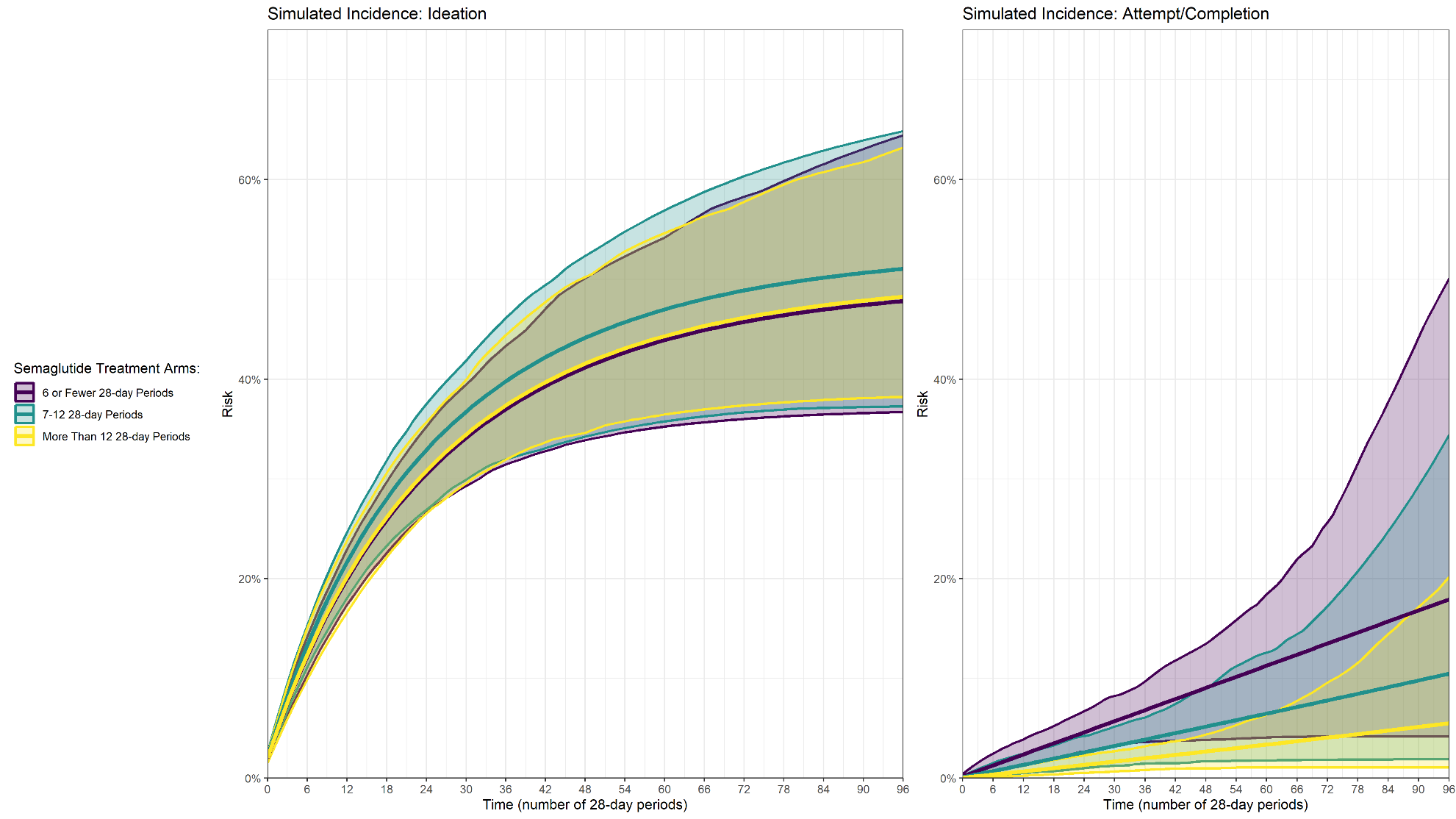
Figure S6. Risk Measure Clone Censor Weighting Simulation Results Among Patients With a Prior History of Suicidality

Figure Legend: Simulated average risk curves depicting the estimated cumulative incidence of suicide ideation (left) and suicide attempt/completion (right) under three hypothetical semaglutide treatment duration strategies among the subgroup of semaglutide initiators with a prior history of suicidality: short-term adherence (Arm A: ≤6 twenty-eight-day periods [approximately ≤6 months]; purple), intermediate adherence (Arm B: 7–12 twenty-eight-day periods [approximately 6–12 months]; teal), and long-term adherence (Arm C: >12 twenty-eight-day periods [approximately >12 months]; yellow). Curves were generated using clone–censor–weight per-protocol methods, with inverse probability of adherence/censoring weights applied to adjust for selection bias introduced by artificial censoring. Shaded bands represent bootstrap-derived 95% confidence intervals based on 1,000 resampled iterations. The x-axis denotes time in twenty-eight-day periods (range 0–96, corresponding to approximately 7.4 years of follow-up). Consistent with the elevated baseline risk in this subgroup, cumulative incidence was substantially higher than in the overall cohort and the no-prior-history subgroup (Figure S5), with suicidal ideation reaching approximately 35–65% and suicide attempt/completion approaching 5–60% by the end of follow-up depending on treatment arm. For suicidal ideation, all three treatment arms followed similar trajectories with broadly overlapping confidence bands, suggesting no significant duration-dependent effect in this subgroup. For suicide attempt/completion, a pronounced dose-response pattern was observed: the short-term adherence arm (Arm A, purple) demonstrated the highest simulated cumulative risk, with marked separation from the long-term adherence arm (Arm C, yellow), which remained the lowest throughout follow-up. The intermediate arm (Arm B, teal) fell between the two. Confidence bands widened considerably at later time periods, reflecting the diminishing number of individuals contributing data at extended follow-up, particularly for the rare outcome of suicide attempt/completion. These findings are consistent with the primary per-protocol analysis (Figure 2), suggesting that the protective association of sustained semaglutide adherence with lower risk of suicide attempts or completions persists among veterans with prior suicidality. Abbreviations: Arm A, ≤6 months semaglutide adherence; Arm B, 6–12 months; Arm C, >12 months.
